# In-silico functional analyses identify *TMPRSS15*-mediated intestinal absorption of lithium as a modulator of lithium response in bipolar disorder

**DOI:** 10.1101/2024.03.27.24304993

**Authors:** David Stacey, Vijayaprakash Suppiah, Beben Benyamin, S Hong Lee, Elina Hyppönen

## Abstract

**Background:** The therapeutic response to lithium in patients with bipolar disorder is highly variable and has a polygenic basis. Genome-wide association studies investigating lithium response have identified several relevant loci, though the precise mechanisms driving these associations are poorly understood. We aimed to prioritise the most likely effector gene and determine the mechanisms underlying an intergenic lithium response locus on chromosome 21 identified by the International Consortium of Lithium Genetics (ConLi^+^Gen).

**Methods:** We conducted in-silico functional analyses by integrating and synthesising information from several publicly available functional genetic datasets and databases including the Genotype-Tissue Expression (GTEx) project and HaploReg.

**Results:** The findings from this study highlighted *TMPRSS15* as the most likely effector gene at the ConLi^+^Gen lithium response locus. *TMPRSS15* encodes enterokinase, a gastrointestinal enzyme responsible for converting trypsinogen into trypsin and thus aiding digestion. Convergent findings from gene-based lookups in human and mouse databases as well as co-expression network analyses of small intestinal RNA-seq data (GTEx) implicated *TMPRSS15* in the regulation of intestinal nutrient absorption, including ions like sodium and potassium, which may extend to lithium.

**Limitations:** Although the findings from this study indicated that *TMPRSS15* was the most likely effector gene at the ConLi^+^Gen lithium response locus, the evidence was circumstantial. Thus, the conclusions from this study need to be validated in appropriately designed wet-lab studies.

**Conclusions:** The findings from this study are consistent with a model whereby *TMPRSS15* impacts the efficacy of lithium treatment in patients with bipolar disorder by modulating intestinal lithium absorption.

## 1. Introduction

Lithium is the gold-standard mood stabilizer for the treatment of bipolar disorder (BPD) (Malhi et al., 2012; Miura et al., 2014). However, the therapeutic response to lithium is highly variable, with ∼30% of patients typically classified as excellent responders, while up to 40% either do not respond at all or experience serious adverse effects including bradycardia, thyroid suppression, and renal dysfunction (Baldessarini and Tondo, 2000; Garnham et al., 2007; Rybakowski et al., 2001). Thus, an improved understanding of the factors driving clinical responses to lithium may help to usher in a precision medicine approach to lithium prescribing.

Studies have shown that clinical predictors of response to lithium do exist, albeit with low sensitivity. These include age of onset, number of hospitalizations, pattern of symptomology, and co-administered medications such as diuretics and antibiotics, which affect lithium kinetics by reducing renal clearance and intestinal absorption, respectively (Kleindienst et al., 2005; Malhi et al., 2020; Rybakowski, 2014). Moreover, a favourable lithium response was more likely in patients with a family history of BPD, and with relatives who have previously responded well (Duffy et al., 2007; Grof et al., 2002; Mendlewicz et al., 1978). The latter is indicative of a role for genetic factors in lithium response.

Although candidate gene approaches to identify genetic factors have largely produced inconsistent results (Senner et al., 2021), genome-wide association studies (GWAS) of lithium response have identified several potentially relevant genetic loci. These include the glutamate decarboxylase like 1 (*GADL1*) (Chen et al., 2014) and SEC 14 and spectrin domain containing (*SESTD1*) (Song et al., 2016) loci, which have been proposed to exert their effects through the regulation of renal lithium clearance (Birnbaum et al., 2014) and phospholipid signalling (Song et al., 2016), respectively. Additionally, a GWAS meta-analysis by the international consortium on Lithium genetics (ConLi^+^Gen) highlighted an intergenic locus on chromosome 21 (Hou et al., 2016), for which the underlying mechanisms have been unclear. Thus, in this study, we sought to characterise this locus by leveraging publicly available online resources and datasets to: (i) prioritise the most likely effector gene, and (ii) elucidate the relevant downstream mechanisms modulating lithium response.

## 2. Methods

### 2.1. Effector gene prioritisation

To avoid confusion with nomenclature from causal inference and mediation analysis methods, we opted to use the term ‘effector gene’ rather than ‘causal’ or ‘mediating’ gene. We employed variant-to-gene (V2G) and gene-to-phenotype (G2P) approaches (**Figure 1A**) to prioritise the most likely effector gene at the ConLi^+^Gen lithium response locus. Briefly, V2G involves searching for functional links between genotype and a locally encoded gene (e.g., genotypic effects on protein structure or local mRNA expression). Conversely, G2P involves searching for evidence of biological links between locally encoded genes and the phenotype of interest or relevant intermediate phenotypes (e.g., genes assigned to phenotypically relevant gene ontology terms or pathways).

**Figure 1.**
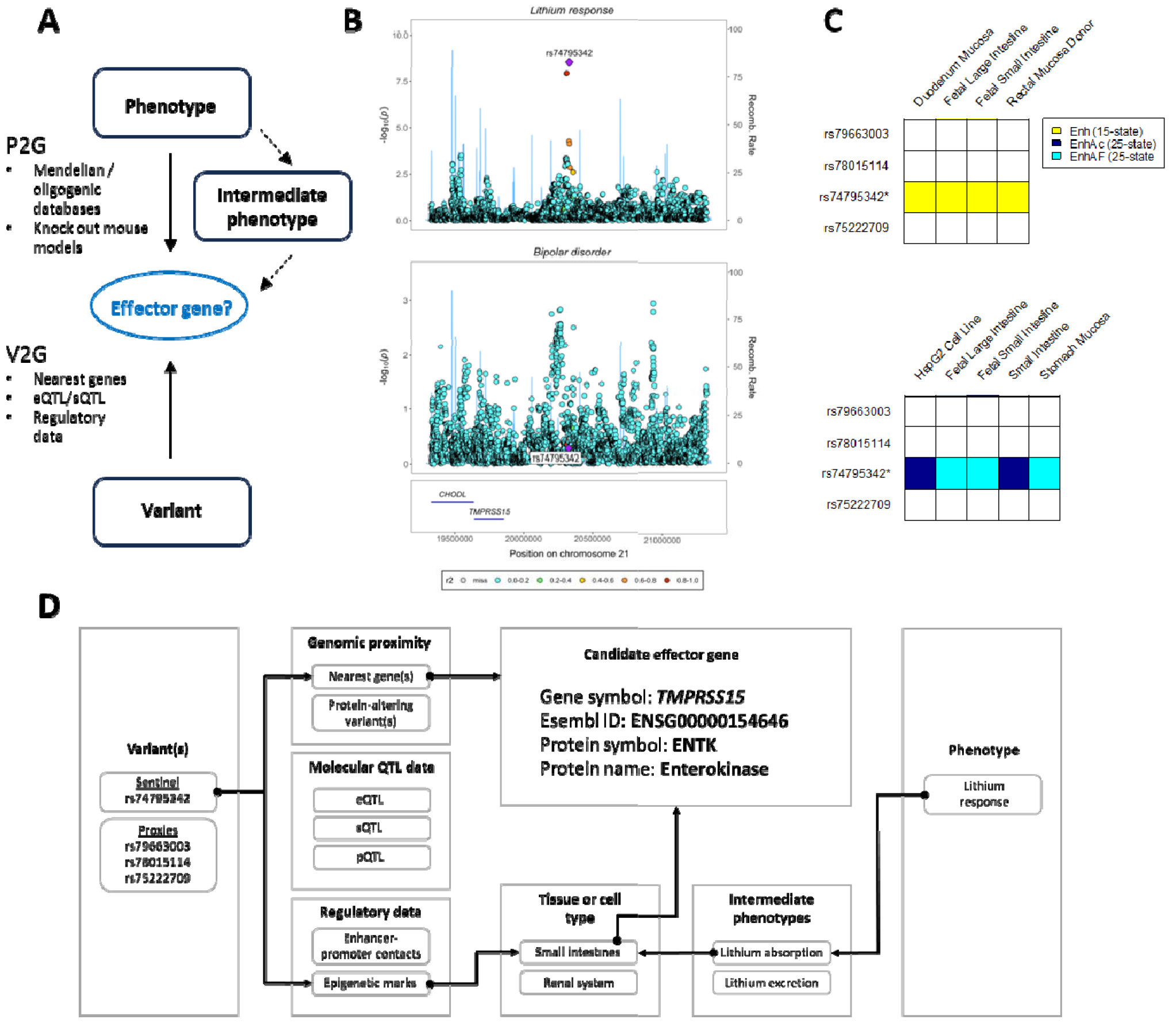
Evidence from public databases converges on *TMPRSS15* as the most likely candidate effector gene at the ConLi^+^Gen lithium response locus. **(A)** Schematic overview of the ‘variant to gene’ (V2G) and ‘phenotype to gene’ (P2G) approaches to prioritising effector genes at disease- or trait-associated loci. **(B)** Stacked regional association plot showing that the ConLi^+^Gen lithium response locus is not a risk locus for bipolar disorder. **(C)** Heatmap showing overlap between lithium response-associated variants at the ConLi^+^Gen locus with gastrointestinal-specific enhancer elements. **(D)** Schematic summarising the lines of evidence converging on *TMPRSS15* as the most likely effector gene at the locus.

For the V2G component, we assessed the sentinel and proxy (r^2^>0.5) variants at the ConLi^+^Gen lithium response locus using the following tools and datasets: (i) the variant effect predictor (VEP) (McLaren et al., 2016); (ii) *cis-*expression and -splice quantitative trait locus (eQTL/sQTL) data from the eQTLgen consortium (whole blood) (Võsa et al., 2021), the Genotype-Tissue Expression (GTEx) project (49 tissues) (GTExconsortium, 2013), and the eQTL catalogue (mixture of tissues and cell types from 30 published studies) (Kerimov et al., 2021); (iii) *cis*-protein QTL (pQTL) data from several published studies accessed through the Open Targets Genetics database (Ghoussaini et al., 2021); and (iv) regulatory data from HaploReg (Ward and Kellis, 2016).

For the G2P searches, since the ConLi^+^Gen locus resides within a gene desert, we selected the three nearest protein-coding genes to the sentinel variant: (i) transmembrane serine protease 15 (*TMPRSS15* [+550 kilobases (kb)]), (ii) chondrolectin (*CHODL* [+680 kb]), and (iii) *C21orf91* [+1.13 megabases (mb)]. We utilised the following databases for the G2P component: (i) the GTEx project (GTExconsortium, 2013) and the human protein atlas (HPA) (Uhlén et al., 2015); (ii) the Online Mendelian Inheritance in Man (OMIM) (Amberger et al., 2014) and Orphanet databases (https://www.orpha.net/consor/cgi-bin/index.php); and (iii) the Mouse Genome Informatics (MGI) database (https://www.informatics.jax.org/).

### 2.2. Co-expression network reconstruction

We reconstructed a co-expression network utilising small intestine read count data (*n*=185) from the GTEx project (v8) (GTExconsortium, 2013). We pre-processed the count data using the edgeR (v3.40.2) R package (Robinson et al., 2010). To remove low abundance genes, we filtered out genes with read counts <10 in >30% samples. We then recalculated library sizes, normalised read counts to counts per million (cpm), and applied a log transformation. To remove the effects of unwanted technical covariates (i.e., recruitment centre, total ischemic time, autolysis time, time tissue spent in PAXgene fixative, batch, and RNA integrity number), we applied empirical bayes-moderate linear regression using the empiricalBayesLM() function from the weighted gene co-expression network analysis (WGNA, v1.72-1) R package (Langfelder and Horvath, 2008). We then reconstructed a signed co-expression network using the WGCNA R package (Langfelder and Horvath, 2008) following procedures described previously (Stacey et al., 2018), with a soft threshold (power) of 12 and minimum cluster size set to 30. We defined the nearest neighbours to *TMPRSS15* as the 200 most correlated genes based on pairwise and absolute Pearson’s correlation coefficients.

### 2.3. Enrichment analyses

We performed gene ontology (GO) term enrichment and pathway analyses using the clusterProfiler (v4.6.2) R package (Yu et al., 2012). In GO term analyses, we examined all three ontological domains: biological process (BP), molecular function (MF), and cellular component (CC) (Ashburner et al., 2000). Pathway analyses were conducted based on the protocol curated by the Kyoto Encyclopedia of Genes and Genomes (KEGG) database (Kanehisa and Goto, 2000). Given that the clusterProfiler package requires entrez gene IDs as input, prior to performing enrichment analyses, we converted ensembl gene IDs into entrez gene IDs using the org.Hs.eg.dg (v3.16.0) R package.

## 3. Results

The sentinel (rs74795342) variant identified by ConLi^+^Gen (*p*<5x10^−8^) (Hou et al., 2016) and all three known proxies (1000G EUR, r^2^>0.8), none of which were associated with risk of BPD (**Figure 1A**) (Mullins et al., 2021), were located within an intergenic gene-poor region on chromosome 21 (q21.1) (**Table S1, Figure 1B**) and is populated by several long non-coding (lnc) RNA genes. The three nearest protein-coding genes, *TMPRSS15, CHODL*, and *C21orf91*, reside more than 550kb, 680kb, and 1.13mb upstream of the sentinel variant, respectively. We annotated additional proxy variants (r^2^>0.5) using the Variant Effect Predictor (VEP) (McLaren et al., 2016), all of which were either intergenic or intronic to a local lncRNA (**Table S1**).

Previous studies have demonstrated that high confidence effector gene predictions can be achieved by integrating several complementary functional genetic datasets (Arnold et al., 2015; Nasser et al., 2021; Stacey et al., 2019). To determine the most likely effector gene at this locus, we initially explored functional links between the lithium response-associated sentinel variant and nearby encoded genes (V2G). We performed an exhaustive search of publicly available human *cis*-expression, -splice, and -protein quantitative trait locus (eQTL/sQTL/pQTL) data (see **Methods**). This search was hampered by missing data due in large part to the remoteness of the locus (i.e., in most cases, the *cis* regions tested around the genes of interest were not large enough to encompass the lithium response-associated variants) and absence of nearby gene features from the summary data (likely due to low abundance). Moreover, of the QTL data that were informative, we found no genotypic associations with *p*<0.01 for any local genes or proteins (**Table S2**).

We then intersected rs74795342 and its proxies (r^2^>0.8) with regulatory data accessible via HaploReg (v4.2) (Ward and Kellis, 2016). We utilised the 15- and 25-state chromatin segmentation model tracks, which were trained on 127 epigenomes from the Roadmap Epigenomes project using 5 and 12 marks, respectively. Both models indicated that the sentinel (rs74795342) variant resided at gastrointestinal-specific enhancers, including fetal small and large intestine (**Figure 1C**). This suggests the locus may only exert regulatory effects in intestinal tissue during early developmental timepoints.

Next, we performed a series of gene-based lookups to identify potential functional links between locally encoded genes and lithium response or relevant intermediate phenotypes (G2P). We first assessed local gene expression profiles using data from the GTEx (GTExconsortium, 2013) and HPA (Uhlén et al., 2015) projects. According to these databases, both *CHODL* and *C21orf91* were expressed ubiquitously across tissues, while *TMPRSS15* expression was tissue-specific and predominantly enriched in the gastrointestinal tract, particularly in the small intestine (**Figure S1**).

Gene-based lookups in the OMIM and Orphanet databases indicated an association between rare loss of function variants affecting *TMPRSS15* and failure to thrive during infancy caused by enterokinase (ENTK) deficiency – the enzyme encoded by *TMPRSS15*. In addition to failure to thrive, ENTK deficiency was characterised by several related symptoms including diarrhea, edema (tissue swelling due to fluid accumulation), and hypoproteinemia (reduced blood protein levels) (Hadorn et al., 1975; Holzinger et al., 2002). Conversely, we found no evidence of a link between *CHODL* or *C21orf91* and any Mendelian or oligogenic phenotypes.

We then complemented our human gene-based lookups by extracting all MPO terms associated with orthologous genes from the MGI database (https://www.informatics.jax.org/). Notably, *Tmprss15*^*-/-*^ mice were annotated to several terms consistent with the failure to thrive phenotype observed in humans, including lower body weight, circulating cholesterol, and total body fat relative to their wild type counterparts (**Table S3**). Furthermore, this mouse model also had elevated circulating levels of sodium and chloride ions. Taken together, the V2G and G2P approaches converge on *TMPRSS15* as a compelling candidate effector gene at the ConLi^+^Gen lithium response locus (**Figure 1D**), while suggesting that the impact of *TMPRSS15* on lithium response may be mediated by altered intestinal lithium absorption (**Figure 1D**).

To validate and further elucidate the proposed role of *TMPRSS15* in modulating lithium absorption, we employed a ‘guilt by association’ approach. We reconstructed a weighted gene co-expression network based on gene expression data from small intestinal tissue of 187 donors from the GTEx project (see **Methods**). Overall, the reconstructed network comprised 10 clusters, with one cluster denoted ‘turquoise’ containing *TMPRSS15* and 4,474 co-expressed genes (**Table S4**).

To characterise the *TMPRSS15*-containing cluster, we conducted gene ontology (GO) term and pathway enrichment analyses using the entire network as a background gene set. Broadly, the enriched terms and pathways were consistent with a role for this cluster in the digestion and metabolism of various metabolites including lipids and proteins, as well as a role in regulating the transport of sodium, chloride, and potassium ions (**Table S5**). For example, we observed an enrichment of genes annotated to ‘lipid catabolic process’ (fold enrichment [FE]=1.88, *p*=1.55x10^−16^), ‘active ion transmembrane transport activity’ (FE=1.83, *p*=7.57x10^−12^), and ‘digestion’ (FE=2.06, *p*=1.70x10^−9^).

Since the *TMPRSS15*-containing cluster was so large (4,474 genes), we created a sub-cluster by extracting the 200 nearest neighbours to *TMPRSS15* (**see Methods**). We again conducted enrichment analyses but this time we used the turquoise cluster as a background gene set. Generally, the findings (**Figures 2B-E, Table S6**) recapitulated those from the previous enrichment analyses. However, in the nearest neighbour analysis we observed stronger enrichment of cellular component GO terms including ‘brush border’ (FE=9.15, *p*=1.42x10^−20^), ‘apical part of cell’ (FE=4.29, *p*=2.93x10^−14^), and ‘microvillus organization’ (FE=10.06, *p*=2.91x10^−7^) (**Figure 2D, Table S6**). Being key components of intestinal enterocytes, these cellular components play central roles in nutrient absorption (Azman et al., 2022). Thus, taken together, these findings are consistent with a role for *TMPRSS15* in regulating intestinal absorption, including ions akin to lithium, like sodium and potassium.

**Figure 2.**
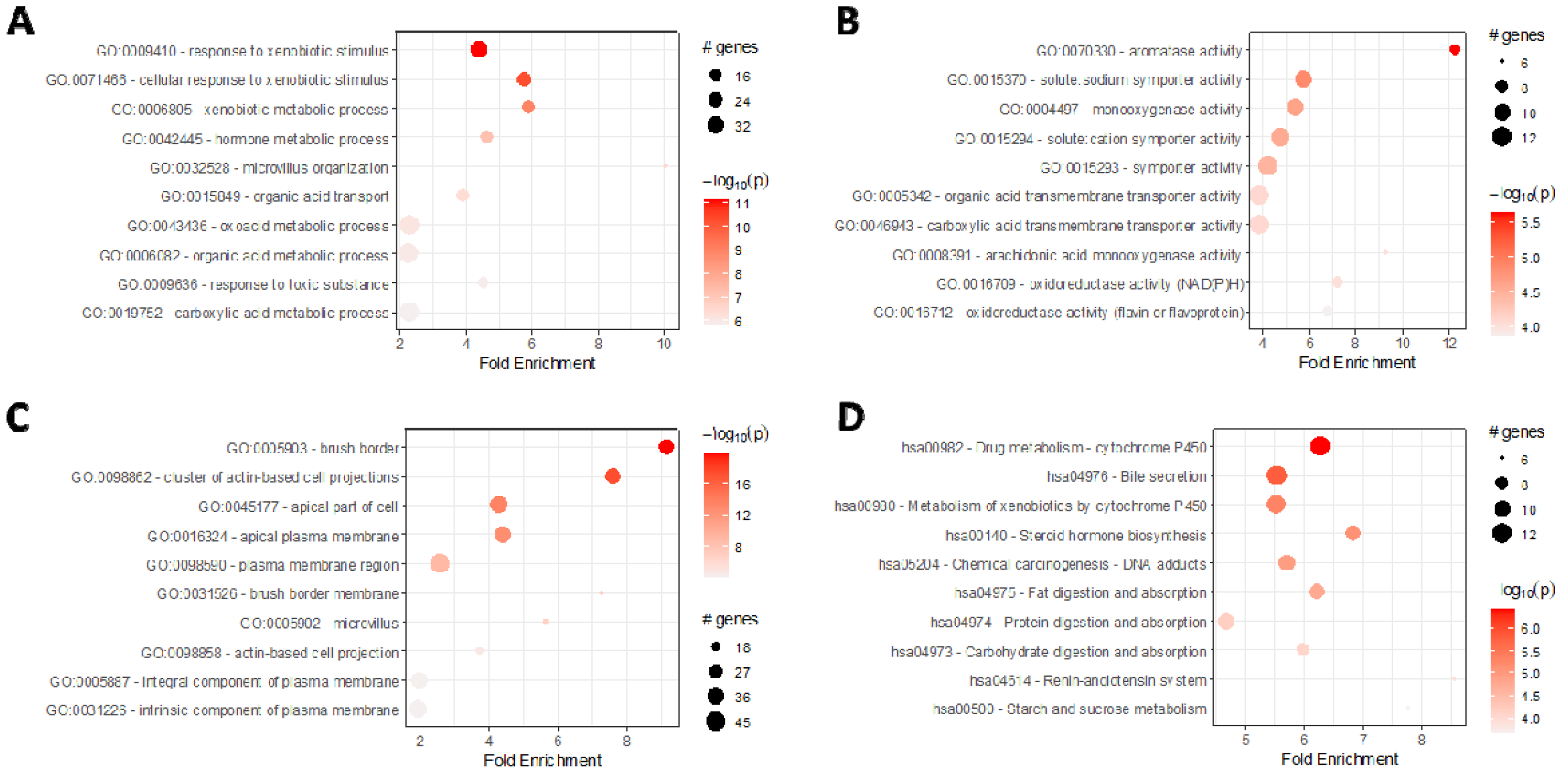
The *TMPRSS15*-containing co-expression cluster is enriched with genes annotated with functional terms and pathways related to intestinal absorption of nutrients, including ion transport. Bubble plots summarising enrichment analyses of the 200 nearest neighbours to *TMPRSS15* using gene ontology (GO) terms from the **(A)** Biological Process, **(B)** Molecular Function, and **(C)** Cellular Component domains, as well as **(D)** Kyoto Encyclopedia of Genes and Genomes (KEGG) pathways.

## 4. Discussion

Here, we sought to functionally characterise an intergenic locus on chromosome 21 implicated in the regulation of lithium response by a GWAS meta-analysis of patients with BPD conducted by ConLi^+^Gen (Hou et al., 2016). Due to the intergenic nature of this locus, the relevant underlying mechanisms have remained elusive, though several previous studies and review articles have suggested that local lncRNAs may be responsible (Hou et al., 2016; Papiol et al., 2022; Senner et al., 2021). However, the findings from this in-silico study indicate that *TMPRSS15*, the nearest protein-coding gene, is the most likely effector gene at this locus.

*TMPRSS15* encodes enterokinase (ENTK), which is localised specifically to gastrointestinal tissues and is best known for its role in aiding digestion by catalysing the conversion of trypsinogen to trypsin (Holzinger et al., 2002). Gene-based lookups and co-expression network analyses converged on a role for *TMPRSS15* in nutrient absorption, including ions such as sodium, potassium, and chloride. Moreover, a recent case study of an enterokinase deficient child found that levels of circulating sodium and potassium ions were below the normal range (Wang et al., 2020). Taken together, we propose that *TMPRSS15* may impact lithium response by modulating the intestinal absorption of lithium, thereby impacting its therapeutic effect.

The genomics of lithium kinetics has not been well studied, with only a single GWAS of plasma lithium levels having been conducted to date. This study consisted of 2,190 Swedish patients with BPD and highlighted a single genome-wide significant locus on chromosome 11. Although the *TMPRSS15* locus was absent from the sub-threshold associations reported in this Swedish study (Millischer et al., 2022), knockdown of *TMPRSS15* in an induced pluripotent stem cell-derived enterocyte monolayer model would constitute a more direct approach to test whether this gene is involved in intestinal lithium transport.

Since lithium has a narrow therapeutic range (between 0.6-1.2 mEq/L) (Nolen and Weisler, 2013), lithium prescribing in patients with BPD usually requires titration to account for inter-individual variability in lithium kinetics (Millischer et al., 2022). The Swedish study above demonstrated that age, sex, and co-medications (e.g., diuretics) can be used to predict plasma lithium levels, though they also showed that the addition of common genetic variants did not improve predictions (Millischer et al., 2022). However, rare variants with large effects on key intestinal (e.g., *TMPRSS15*) or renal genes may still prove to be of predictive value. Thus, future studies may add value by targeting patients with BPD exhibiting abnormally high or low plasma lithium levels – after a comprehensive search for other clinical explanations – for whole exome/genome sequencing.

## 5. Limitations

Although the findings from this study indicate that *TMPRSS15* is the most likely effector gene at the ConLi^+^Gen lithium response locus on chromosome 21, the evidence is circumstantial. Nevertheless, this in-silico study has generated clear and testable hypotheses regarding the role of TMPRSS15 in lithium response, warranting further experimental validation in wet-lab studies.

## 6. Conclusion

The findings from this study are consistent with a model whereby *TMPRSS15* impacts the efficacy of lithium treatment in patients with bipolar disorder by regulating the absorption of lithium in the intestines.

## Data Availability

All data produced in the present study are available upon reasonable request to the authors.

## Declarations of interest

none

## Notes

### Competing Interest Statement

The authors have declared no competing interest.

### Funding Statement

This study did not receive any funding

### Author Declarations

All source data were openly available prior to initiating the study. All data were accessed or downloaded from the following sources: UCSC genome browser (https://genome.ucsc.edu/), eQTLgen (https://www.eqtlgen.org/phase1.html), eQTL catalogue (https://www.ebi.ac.uk/eqtl/), GTEx (https://gtexportal.org/home/), HaploReg (https://pubs.broadinstitute.org/mammals/haploreg/haploreg.php), Orphanet (https://www.orpha.net/), OMIM (https://www.omim.org/), and Mouse genome informatics database (https://www.informatics.jax.org/).

