## Supplementary material for "In-silico functional analyses identify *TMPRSS15*-mediated intestinal absorption of lithium as a modulator of lithium response in bipolar disorder": Figure S1

### Slide 1
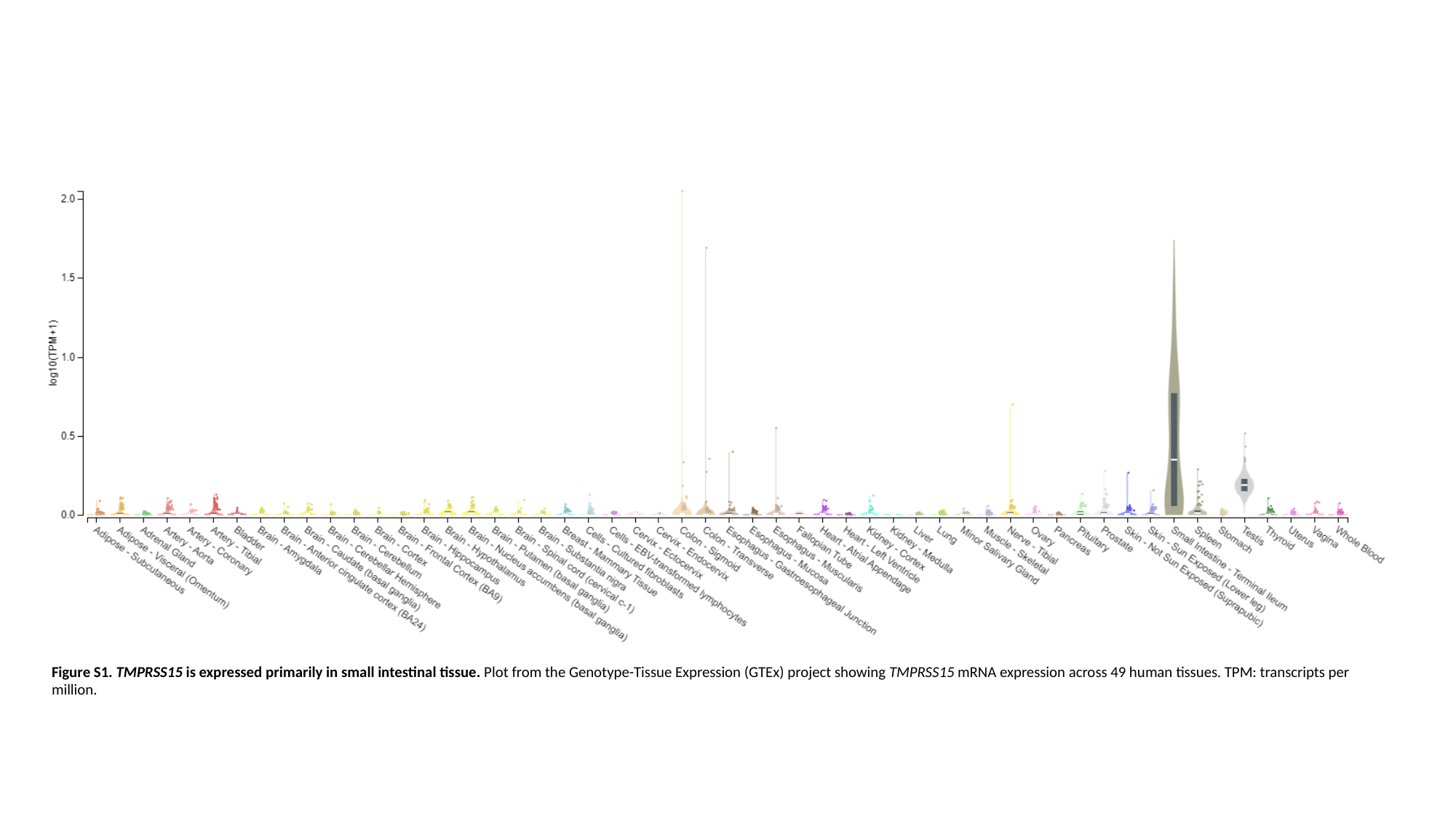

Figure S1. TMPRSS15 is expressed primarily in small intestinal tissue. Plot from the Genotype-Tissue Expression (GTEx) project showing TMPRSS15 mRNA expression across 49 human tissues. TPM: transcripts per million.
